# A multi-ancestry genome-wide association study identifies novel candidate loci in the *RARB* gene associated with hypertensive disorders of pregnancy

**DOI:** 10.1101/2023.10.30.23297806

**Authors:** Jasmine A. Mack, Adam Burkholder, Farida S. Akhtari, John S. House, Ulla Sovio, Gordon C.S. Smith, Charles P. Schmitt, David C. Fargo, Janet E. Hall, Alison A. Motsinger-Reif

## Abstract

**Background:** Genetic factors related to pregnancy-related traits are understudied, especially among ancestrally diverse cohorts. This study assessed maternal contributions to hypertensive disorders of pregnancy (HDP) in multi-ancestry cohorts.

**Methods:** We performed a genome-wide association study of HDP using data from the Personalized Environment and Genes Study (PEGS) cohort (USA) with validation in the UK Biobank (UKBB). We performed gene-level and gene-set analyses and tested the association of polygenic scores (PGS) for systolic blood pressure (SBP), preeclampsia (PE), and gestational hypertension (GH).

**Results:** We identified two novel maternal genome-wide significant associations with HDP. The lead independent variants were rs114954125 on chromosome 2 (near *LRP1B;* OR (95% CI): 3.03 (2.05, 4.49); *P*=3.19 − 10^−8^) and rs61176331 on chromosome 3 (near *RARB;* OR (95% CI): 3.09 (2.11, 4.53); *P*=7.97×10^−9^). We validated rs61176331 in the UKBB (*P=*3.73 − 10^−2^). When aggregating SNPs by genes, *RARB* (*P=*1.36 − 10^−3^) and *RN7SL283P* (*P=*2.56 − 10^−2^) were associated with HDP. Inflammatory and immunological biological pathways were most strongly related to HDP-associated genes. While all blood pressure and HDP-related PGS were significantly associated with HDP in PEGS, the SBP PGS was a stronger predictor of HDP (area under the curve (AUC): 0.57; R^2^=0.7%) compared to the PE PGS (AUC: 0.53; R^2^=0.2%).

**Conclusion:** Our study is the first to identify and validate maternal genetic variants near *RARB* associated with HDP. The findings demonstrate the power of multi-ancestry studies for genetic discovery and highlight the relationship between immune response and HDP and the utility of PGS for risk prediction.

ClinicalTrials.gov Identifier for PEGS: NCT00341237

## Introduction

Hypertensive disorders of pregnancy (HDP) are a heterogeneous set of adverse pregnancy outcomes that occur in the perinatal period comprising gestational hypertension (GH), preeclampsia/eclampsia (PE), chronic, pre-existing hypertension with or without superimposed PE, and postpartum PE. HDP occur in approximately 5-10% of pregnancies worldwide^1^ although prevalence varies by disease subtype and region^2,3^. HDP incidence increased by 11% from 1990 to 2019^4^. Globally, an estimated 50,000-75,000 deaths occur annually due to PE^5^. HDP are the primary cause of maternal morbidity and mortality^2^ in high-resource countries and a secondary cause globally^6^.

There is racial and ethnic disparity regarding HDP prevalence and incidence as well as the contributing risk factors. Black, Hispanic, and Indigenous individuals experience disproportionately high rates of HDP compared to White individuals^7,8^. Risk factors for HDP include pre-pregnancy body mass index (BMI) > 25 kg/m^2^, nulliparity, prior GH, maternal age of more than 35 years, family history of HDP, and pre-existing hypertension^9^.

The pathophysiology of HDP is not fully understood, but dysfunction in placental growth factors and regulators of angiogenesis are suggested mechanisms, with maternal and fetal contributions^10^. This mechanism is hypothesized to be associated with diminished placental perfusion causing systemic vascular endothelial dysfunction through the release of antiangiogenic factors from the placenta^11,12^. Placental hypoperfusion is thought to be caused by failure of transformation of uterine spiral arteries into low-resistance conduits, which is a normal feature of invasion by extravillous cytotrophoblast^13,14^. Failure of spiral artery transformation has been linked to fetal growth restriction, miscarriage, and PE^14^. Moreover, placental factors can interact with maternal genetics, epigenetics, and environmental factors to influence the response to placental dysfunction^15^. Maternal immune system changes are involved in the development and pathogenesis of PE^16–22^, but the mechanisms are incompletely understood.

Early genetic studies of HDP have been predominantly focused on individuals of European ancestry^23,24^. In research in large cohorts with genetic data, participants from underrepresented racial/ethnic groups are consistently excluded from published analyses due to small sample sizes and the use of sparse methods to account for population structure^25–27^.

Prior genome-wide association studies (GWAS) of PE have revealed strong signals near the *FLT1* gene (MIM:165070) based on a meta-analysis of fetal genotypes from five cohorts, of which all participants were of European ancestry: Genetics of Pre-Eclampsia (GOPEC), Avon Longitudinal Study of Parents and Children (ALSPAC), deCODE, Norwegian Mother and Child (MoBa), and Finnish Genetics of Pre-eclampsia Consortium (FINNPEC)^24^. The largest maternal genome-wide multi-ancestry meta-analysis of PE and GH was conducted in cohorts comprising FinnGen, Estonian biobank, Genes and Health, Michigan Genomics Initiative, Mass General Brigham Biobank, Biobank Japan, BioMe Biobank, InterPregGen Consortium, Trøndelag Health Study (HUNT), UKBB, Penn Medicine Biobank, and Nulliparous Pregnancy Outcomes Study: Monitoring Mothers-to-Be (nuMoM2b)^28^. Among 31,091 HDP cases assessed in the meta-analysis, 83% were of European ancestry. Multi-ancestry meta-analyses have been conducted in diverse cohorts such as the Pan UK Biobank (UKBB), but the control groups of the GWAS also included female UKBB participants who have never been pregnant^29^.

To increase diversity and support the representation of non-European populations in genomics research, GWAS that include minority racial and ethnic groups can be conducted in currently available data from diverse cohorts. The results can be stratified by ancestry, and meta-analyses can jointly model effects in a multi-ancestry analysis. Building upon previous multi-ancestry genetic studies, we conducted multi-ancestry GWAS in whole-genome sequencing (WGS) data from a discovery cohort from the North Carolina-based Personalized Environment and Genes Study (PEGS) using a case-control study design. The joint modeling exhibited improved power for genetic discovery, enabling the first discovery of a maternal genetic association near the retinoic acid receptor (*RARB*) gene (MIM: 180220) with HDP. We validated this association in the UKBB and a meta-analysis in PEGS and the UKBB. In downstream analyses, we performed gene-level analysis and gene-set analyses to elucidate the biological mechanisms involved in HDP. Further, in the PEGS dataset, we assessed the predictive ability of existing polygenic scores (PGS) for PE and GH, with HDP in predicting risk of these conditions.

## Methods

The data used in this study are available to researchers through application to dbGAP the UK Biobank (https://www.ukbiobank.ac.uk/register-apply/). Reference panel data from the 1000 Genomes Project and polygenic scores from the PGS Catalog are publicly available. The Major Resources Table provides details. The summary statistics that support the study findings are available from the corresponding author upon reasonable request.

### Study Populations

#### PEGS Cohort

For the discovery cohort, we selected participants from PEGS, a long-term, prospective cohort based in North Carolina^30^. PEGS collects extensive clinical, genetic, environmental, and self-reported health information to investigate disease etiology and risk factors across phenotypes in a diverse group of 19,685 individuals.

Participants are administered the Health and Exposure Survey, External Exposome Survey, and Internal Exposome Survey that request information on sociodemographic factors, individual and family medical history, lifestyle factors, medication use, diet, and physical environmental exposures from childhood to adulthood. While PEGS exclusion criteria include women who self-report being pregnant, a small number of participants were pregnant when they completed the Internal Exposome Study, as this was administered later in the data collection process.

For a subset of 4,737 PEGS participants who provided blood and urine samples, WGS data were processed, with the optional return of results for select health conditions. The Broad Institute sequenced all samples on the NovaSeq 6000 platform with a target genome-wide read depth of 30x (24 samples per flow cell).

For downstream analyses, we removed related individuals in the PEGS genomic cohort by analyzing the genotypes of all individuals using KING 2.2.5^31^ to generate pairwise kinship estimation. We excluded second-degree or closer relatives using a kinship threshold of 0.0884. We estimated genome-wide local ancestry on the autosomes and X chromosome using RFMix version 2, a discriminative modeling approach^32^. We classified genomic positions by five genetic similarity groups: African (AFR), Admixed American (AMR), East Asian (EAS), European (EUR), and South Asian (SAS). We classified individuals into one of the genetic similarity groups if any genome-wide local ancestry proportion was greater than 0.50. Otherwise, we classified participants as “None”.

The PEGS cohort is a registered clinical trial, and all participants provided written informed consent (ClinicalTrials.gov Identifier: NCT00341237)^30^. We obtained the data used for the analyses from PEGS Data Freeze version 2.0 created in August 2021. The **Supplemental Methods** section provides details on genome sequencing, quality control, and local ancestry inference.

#### UK Biobank

For the validation cohort, we selected participants from the UK Biobank (UKBB). UKBB is a large, deeply phenotyped genomic cohort comprising 488,366 participants aged 40-69 years at recruitment from 2006 to 2010. About 6% of participants (>30,000) identified as having a non-White ethnic background^25,33^. Genetic data are linked to electronic health records and survey data, and UKBB has information on diagnosis history, brain imaging, medication use, lifestyle factors, deprivation indices, environmental exposure data, and sociodemographic characteristics.

Ethical approval for UKBB was obtained from the North West Centre for Research Committee (11/NW/0382). Informed consent was obtained for all participants. The following work described was approved by the UKBB team under Application No. 57849. We obtained predominant ancestry assignments from UKBB Returned Dataset 2442^34^. The **Supplemental Methods** provides details on genotyping and genetic similarity groups in UKBB.

##### Case-Control Definition

**Figure S1** outlines the steps taken to define cases and controls in the PEGS cohort. Because the data available for PEGS and UKBB are not standardized, we defined cases and controls separately for each cohort and harmonized the case-control sets as much as possible. The **Supplemental Methods** section provides details on case-control definition.

#### PEGS

We defined cases and controls for 4,933 PEGS participants who self-reported at least one pregnancy. After filtering, there were 202 GH cases, which is the primary outcome, 92 PE cases, 78 GH only cases, 223 GH/PE cases, and 1,569 controls. **Table S1** lists the phenotypes and survey questions used to define cases and controls.

#### UKBB

We defined cases and controls in the UKBB population using the International Classification of Diseases (ICD) codes (ICD10) for pregnancy-induced hypertension with and without significant proteinuria (PE and hemolysis, elevated liver enzymes and low platelets (HELLP) syndrome), eclampsia, and unspecified maternal hypertension (**Table S1**). After filtering, there were 1,319 HDP cases and 2,050 controls.

### Statistical Analysis

For each cohort, we calculated frequencies and proportions for categorical variables and mean/standard deviation for normally distributed continuous variables, including covariates and sociodemographic characteristics. For continuous variables without normal distribution, we report median and interquartile ranges. We estimated *P* values by Fisher’s exact test to assess associations among covariates and performed t-tests for continuous variables. For variables without normal distribution, such as BMI, we used the Mann-Whitney U test.

#### Genome-wide Association Study for Common Variants

We conducted GWAS (MAF>=0.03) using multivariable logistic regression to model the odds of HDP, assuming an additive allelic genetic model. We adjusted for the top 10 genetic principal components, self-reported race/ethnicity, age and BMI at enrollment, and gravidity. We used PLINK2.0^35^ for all GWAS with Firth-corrected logistic regression modeling to account for case-control imbalance^36^. Because information on gravidity was available for few UKBB participants, we excluded this covariate in the UKBB analyses. We included genotyping batch as a covariate in the UKBB analyses given that two types of arrays were used in genotyping with 106 batches^25^. We used a threshold of *P* < 5 × 10^−8^ for genome-wide significance and *P* < 1 × 10^−6^ for suggestive significance.

In addition to the main outcome of HDP, we performed GWAS for three other phenotypes in the PEGS discovery cohort (PE, GH without PE, and GH and/or PE) for all individuals and separately for the AFR and EUR similarity groups. We adjusted for within-group variation using the top 10 principal components, age and BMI at enrollment, and gravidity. The AMR, EAS, and SAS similarity groups had insufficient numbers of participants to perform stratified analyses (**Table 1**).

**Table 1.**
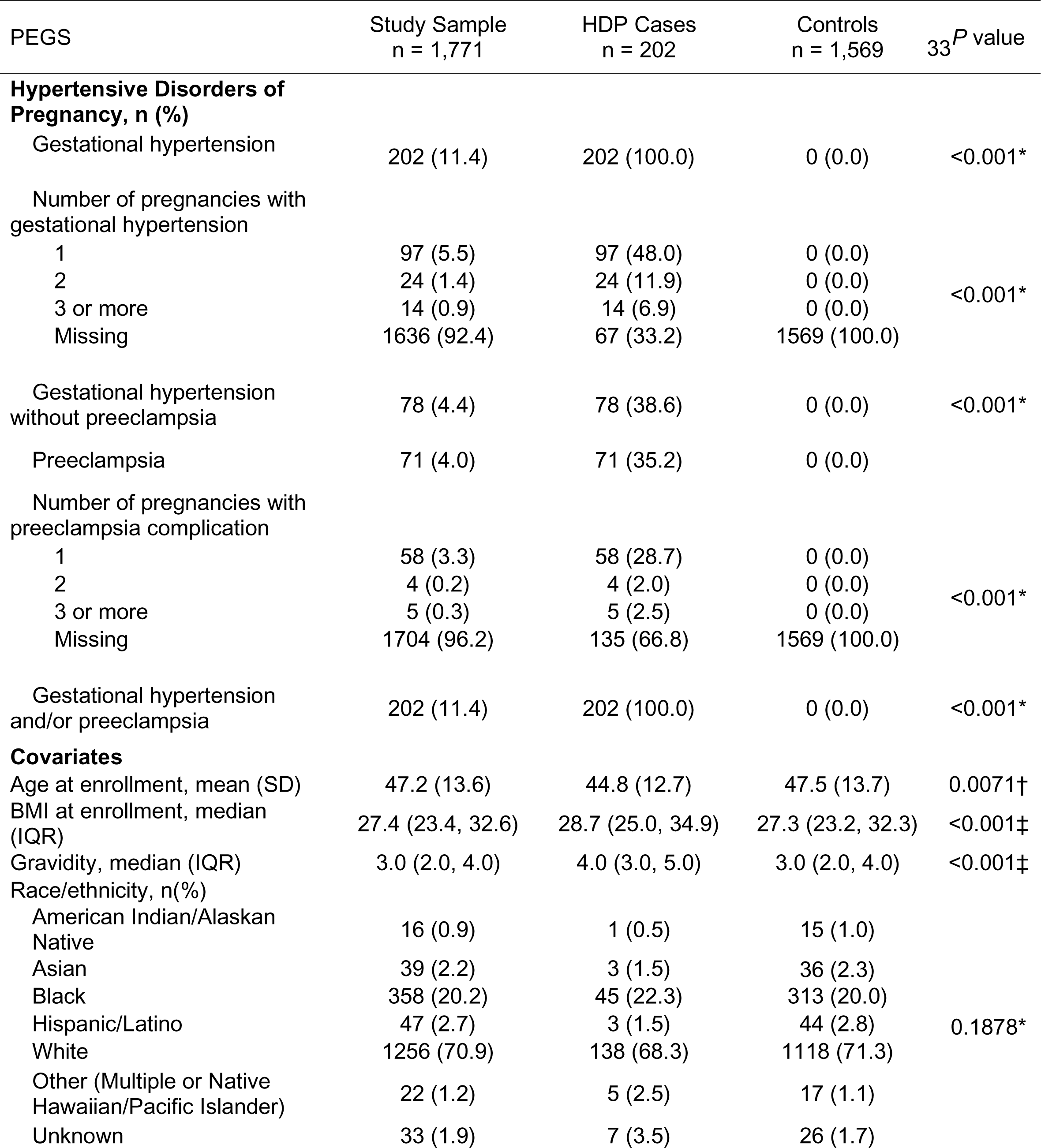
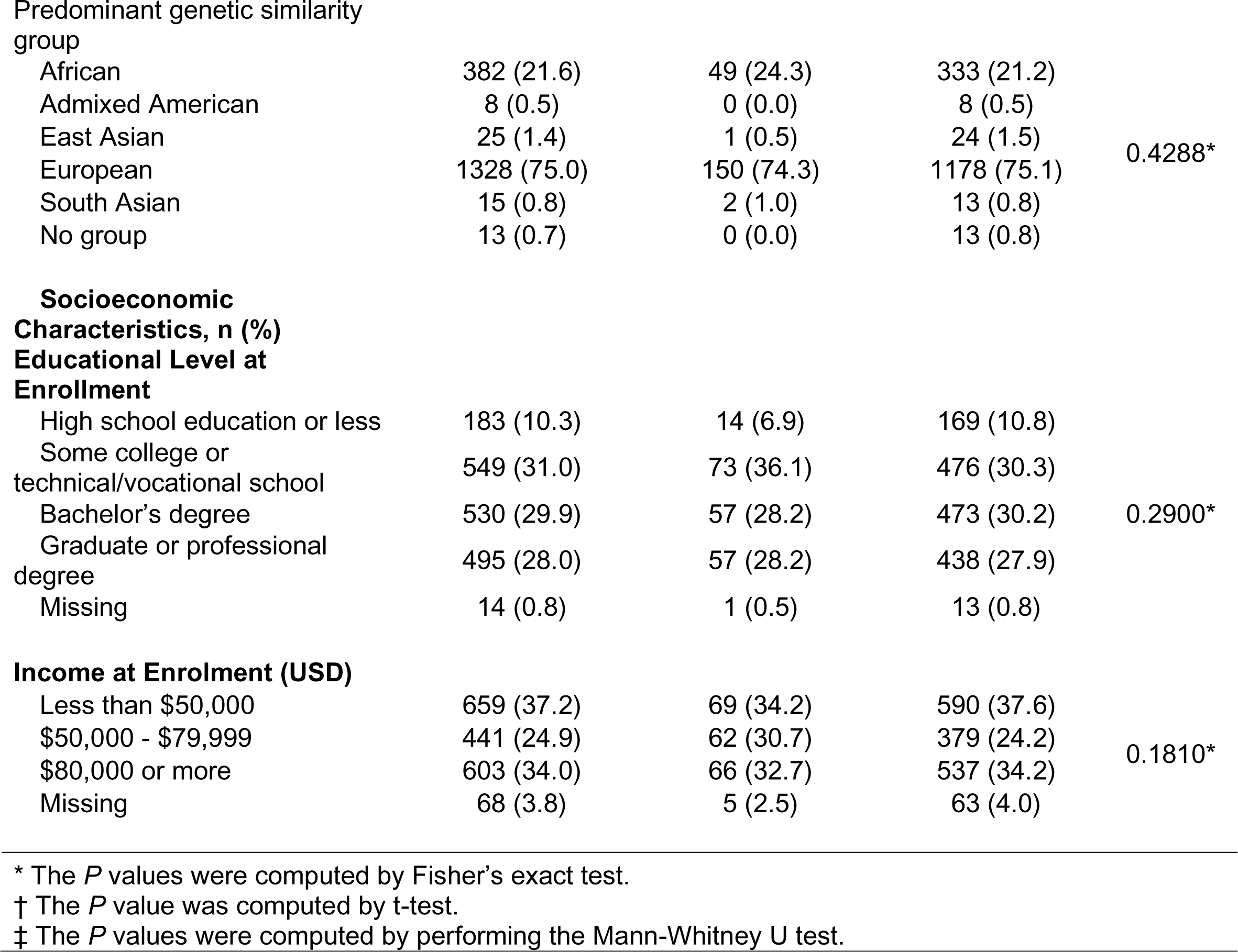
Demographic characteristics of participants in the PEGS discovery cohort.

| PEGS | Study Sample<br>n = 1,771 | HDP Cases<br>n = 202 | Controls<br>n = 1,569 | 33<sup>P</sup> value |
| --- | --- | --- | --- | --- |
| **Hypertensive Disorders of Pregnancy, n (%)** |  |  |  |  |
| Gestational hypertension | 202 (11.4) | 202 (100.0) | 0 (0.0) | <0.001* |
| Number of pregnancies with gestational hypertension |  |  |  |  |
| 1 | 97 (5.5) | 97 (48.0) | 0 (0.0) | <0.001* |
| 2 | 24 (1.4) | 24 (11.9) | 0 (0.0) |  |
| 3 or more | 14 (0.9) | 14 (6.9) | 0 (0.0) |  |
| Missing | 1636 (92.4) | 67 (33.2) | 1569 (100.0) |  |
| Gestational hypertension without preeclampsia | 78 (4.4) | 78 (38.6) | 0 (0.0) | <0.001* |
| Preeclampsia | 71 (4.0) | 71 (35.2) | 0 (0.0) |  |
| Number of pregnancies with preeclampsia complication |  |  |  |  |
| 1 | 58 (3.3) | 58 (28.7) | 0 (0.0) | <0.001* |
| 2 | 4 (0.2) | 4 (2.0) | 0 (0.0) |  |
| 3 or more | 5 (0.3) | 5 (2.5) | 0 (0.0) |  |
| Missing | 1704 (96.2) | 135 (66.8) | 1569 (100.0) |  |
| Gestational hypertension and/or preeclampsia | 202 (11.4) | 202 (100.0) | 0 (0.0) | <0.001* |
| **Covariates** |  |  |  |  |
| Age at enrollment, mean (SD) | 47.2 (13.6) | 44.8 (12.7) | 47.5 (13.7) | 0.0071† |
| BMI at enrollment, median (IQR) | 27.4 (23.4, 32.6) | 28.7 (25.0, 34.9) | 27.3 (23.2, 32.3) | <0.001‡ |
| Gravidity, median (IQR) | 3.0 (2.0, 4.0) | 4.0 (3.0, 5.0) | 3.0 (2.0, 4.0) | <0.001‡ |
| Race/ethnicity, n(%) |  |  |  |  |
| American Indian/Alaskan Native | 16 (0.9) | 1 (0.5) | 15 (1.0) | 0.1878* |
| Asian | 39 (2.2) | 3 (1.5) | 36 (2.3) |  |
| Black | 358 (20.2) | 45 (22.3) | 313 (20.0) |  |
| Hispanic/Latino | 47 (2.7) | 3 (1.5) | 44 (2.8) |  |
| White | 1256 (70.9) | 138 (68.3) | 1118 (71.3) |  |
| Other (Multiple or Native Hawaiian/Pacific Islander) | 22 (1.2) | 5 (2.5) | 17 (1.1) |  |
| Unknown | 33 (1.9) | 7 (3.5) | 26 (1.7) |  |
| African | 382 (21.6) | 49 (24.3) | 333 (21.2) | 0.4288* |
| Admixed American | 8 (0.5) | 0 (0.0) | 8 (0.5) |  |
| East Asian | 25 (1.4) | 1 (0.5) | 24 (1.5) |  |
| European | 1328 (75.0) | 150 (74.3) | 1178 (75.1) |  |
| South Asian | 15 (0.8) | 2 (1.0) | 13 (0.8) |  |
| No group | 13 (0.7) | 0 (0.0) | 13 (0.8) |  |
| High school education or less | 183 (10.3) | 14 (6.9) | 169 (10.8) | 0.2900* |
| Some college or<br>technical/vocational school | 549 (31.0) | 73 (36.1) | 476 (30.3) |  |
| Bachelor's degree | 530 (29.9) | 57 (28.2) | 473 (30.2) |  |
| Graduate or professional<br>degree | 495 (28.0) | 57 (28.2) | 438 (27.9) |  |
| Missing | 14 (0.8) | 1 (0.5) | 13 (0.8) |  |

|  |  |  |  |  |
| --- | --- | --- | --- | --- |
| Less than \$50,000 | 659 (37.2) | 69 (34.2) | 590 (37.6) | 0.1810* |
| \$50,000 - \$79,999 | 441 (24.9) | 62 (30.7) | 379 (24.2) | |
| \$80,000 or more | 603 (34.0) | 66 (32.7) | 537 (34.2) | |
| Missing | 68 (3.8) | 5 (2.5) | 63 (4.0) |  |

\* The *P* values were computed by Fisher's exact test.
† The *P* value was computed by t-test.
‡ The *P* values were computed by performing the Mann-Whitney U test.

To validate our findings, we filtered GWAS summary statistics from the PEGS multi-ancestry analysis to include only variants with suggestive association (*P <* 1 × 10^−6^). We assessed these variants in the UKBB using harmonized cases and controls. We used GCTA-COJO for conditional and joint analyses to identify independently associated loci^37^. We used METAL to perform inverse variance-weighted meta-analyses for variants with suggestive association in PEGS and UKBB^38^.

#### Gene-based Association Analysis and Gene-set Analysis

To increase statistical power for discovery, we used MAGMA to perform gene-set association analysis, adjusting for the top 10 genetic principal components, self-reported race/ethnicity, age and BMI at enrollment, and gravidity. We used Ensembl annotation on GRCh38 to map the SNVs from the GWAS to 31,427 genes^39^. We utilized the results from the gene-level analysis to perform gene-set analysis in MAGMA^40,41^. We used the Kyoto Encyclopedia of Genes and Genomes (KEGG) to define gene sets by biological pathways and conditions^42,43^. To control the false discovery rate (FDR), we used the Benjamini-Hochberg method to adjust *P* values from the statistical analyses for multiple comparisons. We define statistical significance by an adjusted *P* value of 0.05 and suggestive FDR-corrected significance by an adjusted *P* value of 0.10.

### Application of Polygenic Scores

To assess the predictive value of SBP, PE, and GH for HDP in PEGS, we applied an SBP PGS derived from the UKBB female sub-cohort in Kauko et al.^44^ and separate scores for SBP, PE, and GH from meta-analyses in Honigberg et al.^28^. We separately computed the four PGS in the PEGS pregnancy (n = 1,771) and female sub-cohorts (n = 3,102) to model HDP and essential hypertension, respectively. We standardized each PGS. The **Supplemental Methods section** provides details.

We performed logistic regression with Firth correction to assess the association between HDP risk and the standardized PGS, adjusting for the top ten genetic principal components, self-reported race/ethnicity, age and BMI at enrollment, and gravidity. For further validation, we tested the association of each PGS with essential hypertension in the PEGS female sub-cohort, adjusting for the same covariates except gravidity. Additionally, we performed stratified analyses for the AFR and EUR genetic similarity groups. For each PGS, we report odds ratios per standard deviation with a 95% CI. We calculated the AUC for each model using the pROC package in R to assess discriminative ability. All PGS analyses were performed in R 4.3.1 with a significance threshold of 0.05.

The Major Resources Table in the **Supplemental Materials** provides details on data and code availability.

## Results

In the PEGS cohort, participants with HDP (11.4% of the study sample) were younger, had a higher BMI at enrollment, and had more pregnancies compared to participants in the control group (*P*<0.001; **Table 1**). Among HDP cases, 4.5% experienced recurrent diagnoses of PE, and 18.8% experienced GH during two or more pregnancies. Race/ethnicity distribution, education level, and income were similar for the case and control groups (**Table 1**). In the UKBB cohort, race/ethnicity, predominant genetic similarity group, and education level were similar for cases and controls (**Table S2**). Similar to PEGS, UKBB participants with HDP were younger (*P*=0.024) and had a higher BMI at recruitment (*P*<0.001) compared to controls.

### Novel Maternal Loci Identified on *RARB*

In the PEGS GWAS, 11 variants in two loci met genome-wide significance (**Figure 1A; Table S3**) for HDP. An association was found for a significant novel candidate region in the maternal genome (202 cases and 1,569 controls) consisting of intergenic variants near the low-density lipoprotein (LDL) receptor-related protein 1B (*LRP1B;* MIM:608766) gene on chromosome 2 (**Figure 1B**). A stronger signal was observed for a region identified as intronic to the *RARB* gene (MIM:180220) on chromosome 3 (**Figure 1C**). The lead variant was rs61176331, with an OR of 3.09 for HDP (EAF: 0.048; 95% confidence interval (95% CI): (2.11, 4.53); *P=*7.97 − 10^−9^). The second most significant variant was rs114954125, with an OR of 3.03 (EAF: 0.051; 95% CI: (2.05, 4.49); *P=*3.19 − 10^−8^) (**Table 2**). A genomic inflation factor of 1.02 indicated sufficient adjustment for population stratification (**Figure S2A**). Conditional and joint analyses confirmed that rs61176331(*P*_joint_=9.56 − 10^−9^) and rs114954125 (*P*_oint_=3.79 − 10^−8^) are independent loci (**Table 2**). In results stratified by genetic similarity group, these loci had larger effect sizes although the *P* values were attenuated. rs61176331 had an OR of 6.98 and 2.37 for the AFR and EUR genetic similarity groups, respectively (**Table 2; Figure S2B**). rs114954125 had a similar effect size as the multi-ancestry GWAS for the EUR genetic similarity group, but this variant did not reach the quality control threshold in the AFR genetic similarity group. We validated rs61176331 in the UKBB analysis (OR: 1.29; 95% CI: (1.02, 1.65); *P=*3.73 − 10^−2^) (**Figure S2B**). In the meta-analysis including PEGS and UKBB, strong associations remained for the candidate region near *RARB* (**Table 2**; **Table S3**).

**Figure 1.**
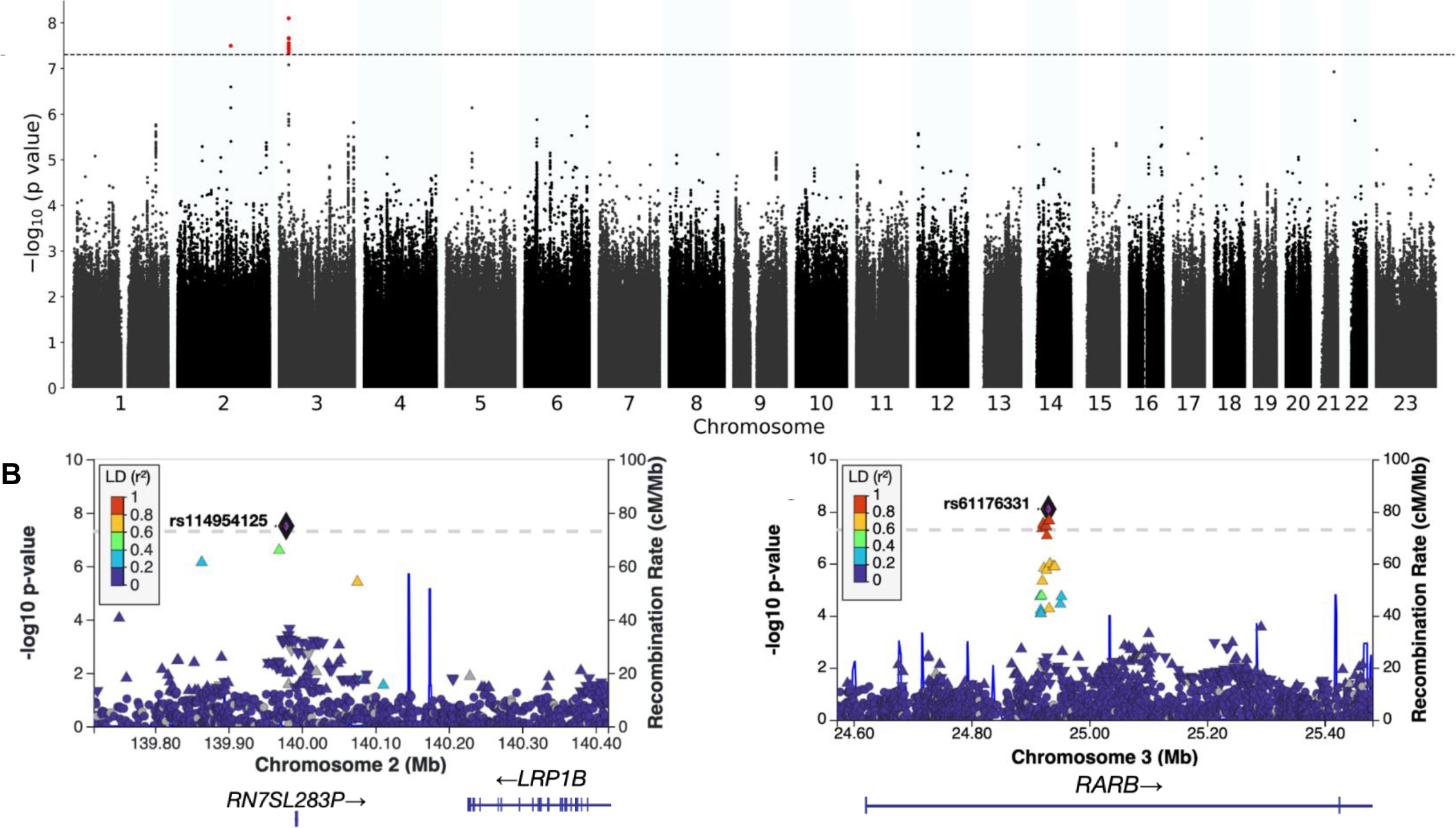
Manhattan plot of hypertensive disorders of pregnancy GWAS results and LocusZoom plots of genome-wide significant loci in the PEGS cohort (202 cases and 1,569 controls). The genome coordinates for **B** and **C** reflect GRCh38 assembly.

**Table 2.**
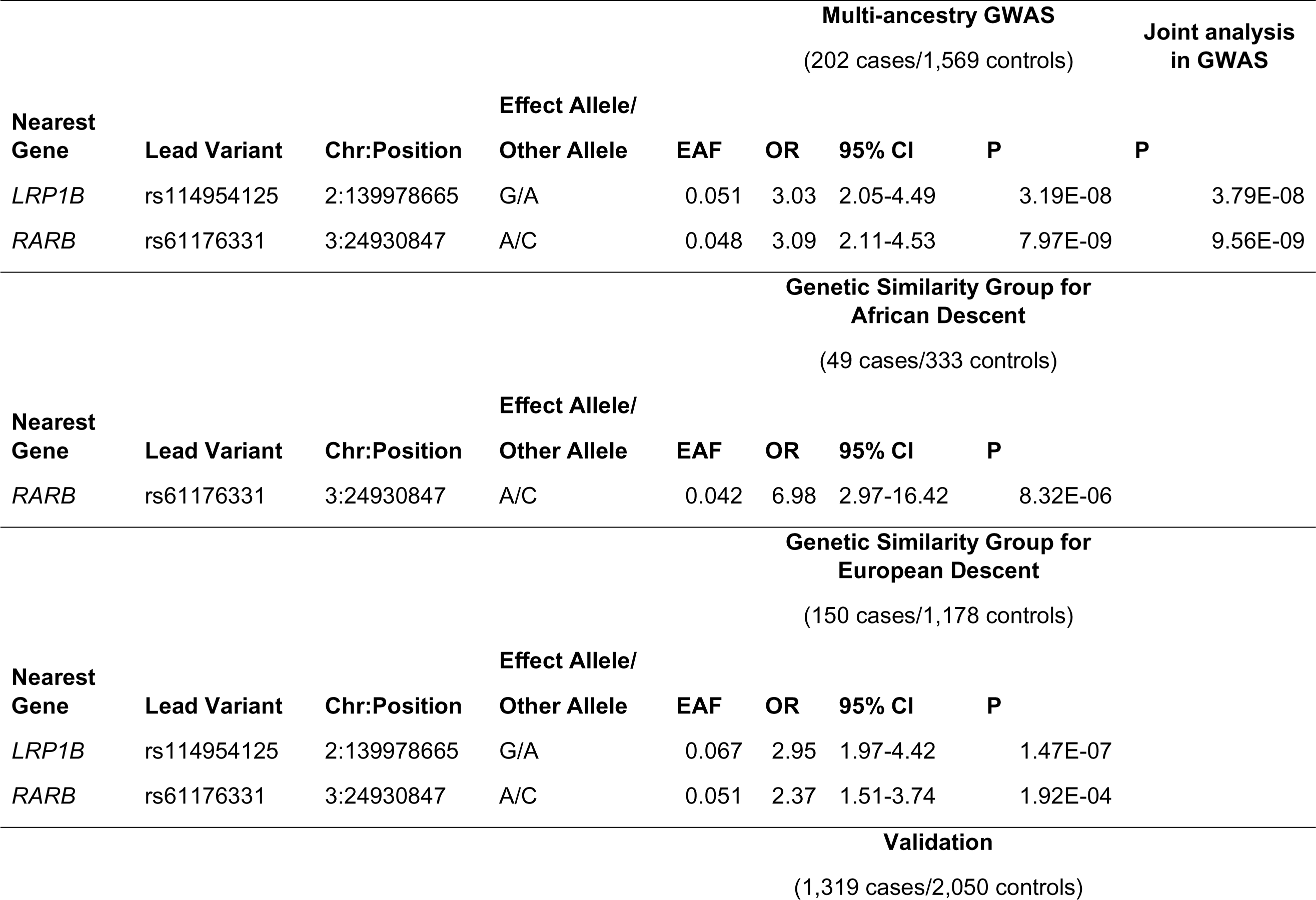
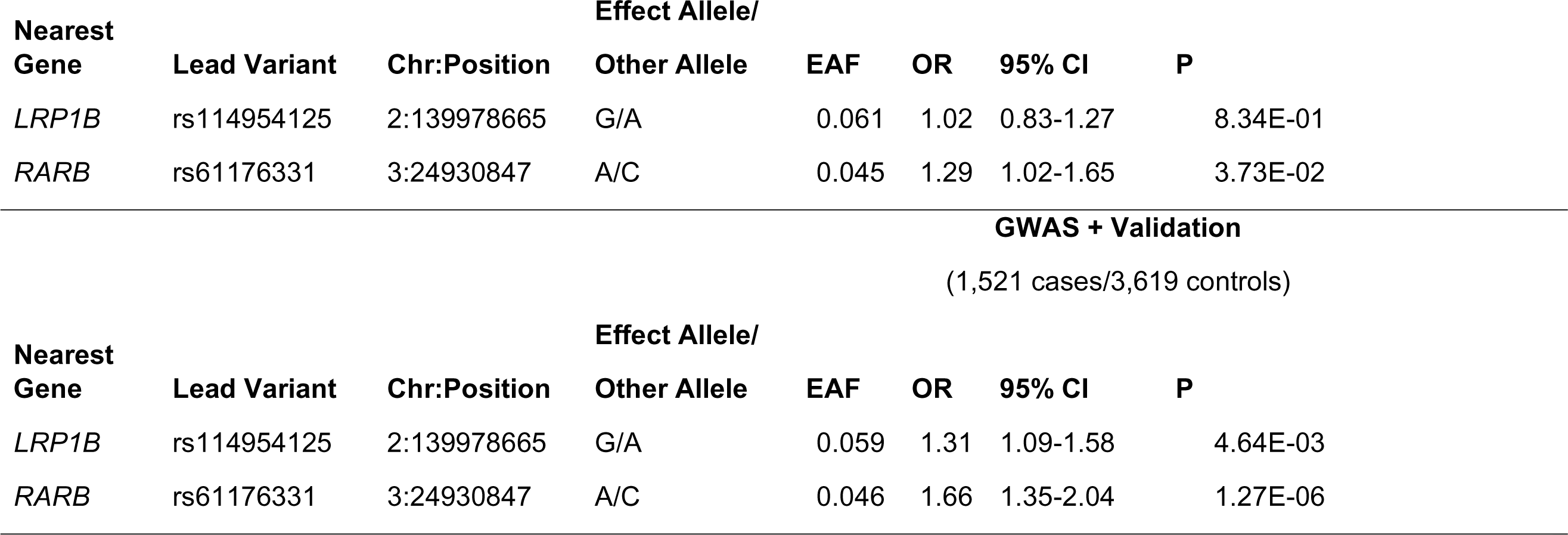
Summary statistics for rs114954125 (*LRP1B*) and rs61176331 (*RARB)*. Chr = chromosome, EAF = effect allele frequency.

In a sensitivity analysis with three HDP subtypes as the outcome (PE, GH only, and GH and/or PE; **Figure S3**), there was distinct attenuation of signals in the multi-ancestry HDP GWAS with PE as the outcome (21 cases) (**Figure S3A**). In this GWAS, the candidate signal on *RARB* was attenuated below the genome-wide significance threshold while rs114954125 on chromosome 2 remained significant. (**Figure S3E**).

### Significant Gene-wide Associations for *RARB*

**Figure 2** is a gene-based Manhattan plot. Two gene signals met the FDR-corrected significance threshold—*RARB* (*P=*1.36 − 10^−3^) and *RN7SL283P (P=*2.56 − 10^−3^). Among the top 15 most significant genes, there were strong peaks on chromosome 6 (*HLA-DRB5, BTNL2, RNU1-61P*) and chromosome 17 (*LRRC59, EME1, ACSF2;* **Table S4**) although these associations did not reach gene-wide significance.

**Figure 2.**
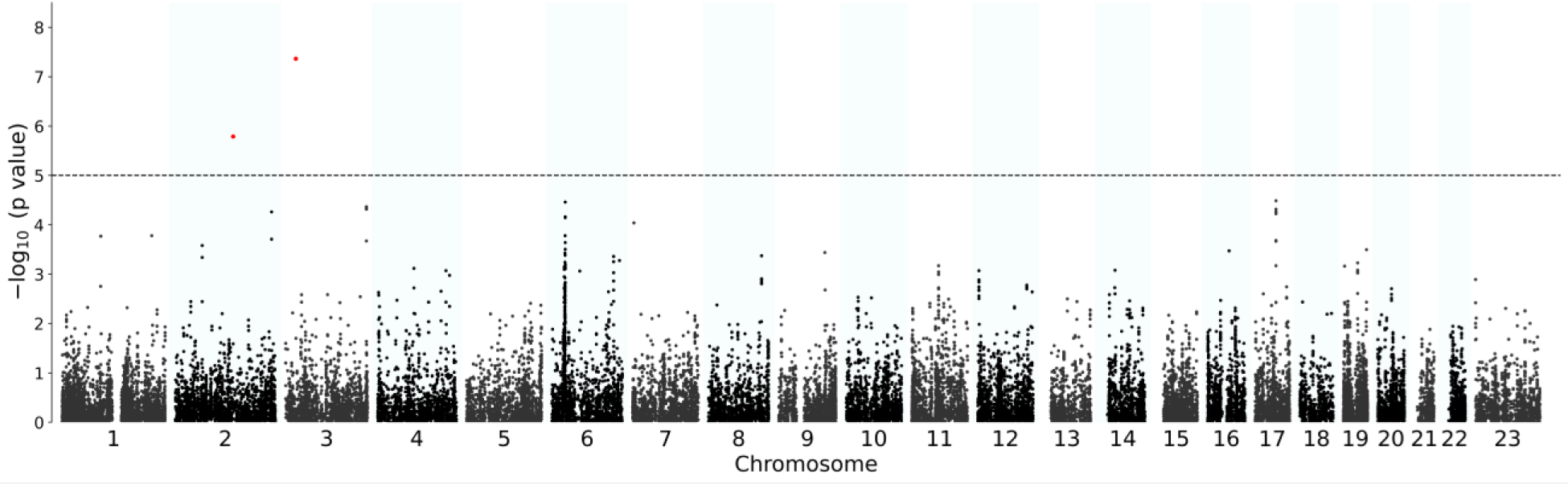
Manhattan plot of gene-based association analysis of hypertensive disorders of pregnancy. Labeled genes represent the nearest gene within 50 kb upstream or downstream of the gene.

### Immune-related Pathways Linked to HDP

Among the ten most significant KEGG gene sets associated with HDP were the pentose phosphate pathway and the hedgehog signaling pathway (**Figure 3**). Gene-set analyses revealed additional immune-related diseases and processes related to HDP-associated genes (**Table S5**), but these sets, which included autoimmune thyroid disease, graft-versus-host disease, adipocytokine signaling pathway, and allograft rejection (uncorrected *P*<0.05), did not meet the FDR-corrected significance threshold.

**Figure 3.**
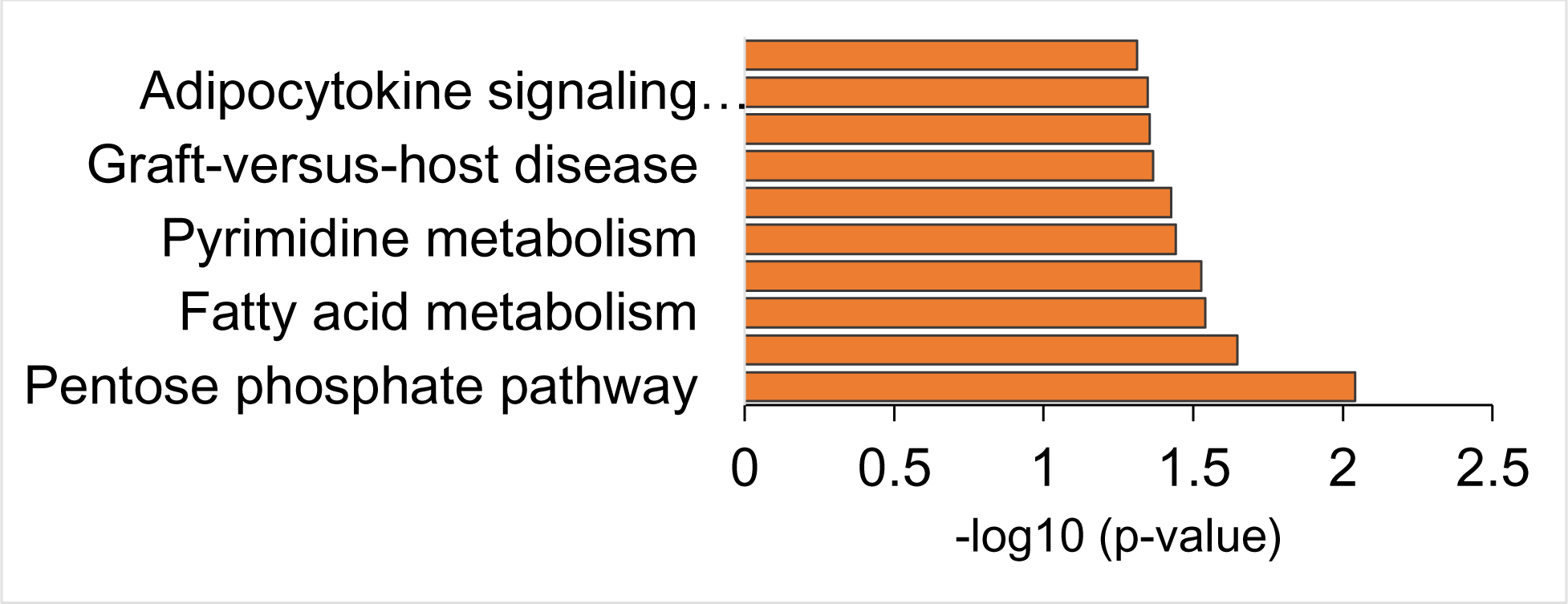
Gene-set analysis using KEGG with MAGMA. The ten most significant KEGG sets are shown on the y-axis. Corresponding unadjusted -log_10_(p-values) are shown on the x-axis. *P* values were estimated by linear regression model-based testing using MAGMA.

### Polygenic Score for Systolic Blood Pressure is Associated with HDP

We tested the association of HDP with PGS for SBP, PE, GH with HDP, and essential hypertension in the PEGS female sub-cohort. **Table S6** shows the number of variants missing in the PEGS cohort for each PGS. In the overall model, all four PGS were significantly associated with HDP (*P*<0.05). The AUC (0.57; 95% CI: (0.52-0.61); **Figure 4; Figure S5**) and pseudo-R^2^ (0.7%; **Table S7**) were the highest for the SBP PGS [1] based on the UKBB. One standard deviation increase in the SBP PGS [1] was associated with a 2.09-fold increase in HDP odds (95% CI: 1.40, 3.13). GH-PGS had the highest OR for HDP (**Figure 4**). **Figure S6** shows HDP prevalence by percentile for each PGS. When the results were stratified by genetic similarity group, the four PGS were not significantly associated with HDP in the AFR genetic similarity group, but there were similar effect sizes for the overall models and for the EUR genetic similarity group (**Figure 4**).

**Figure 4.**
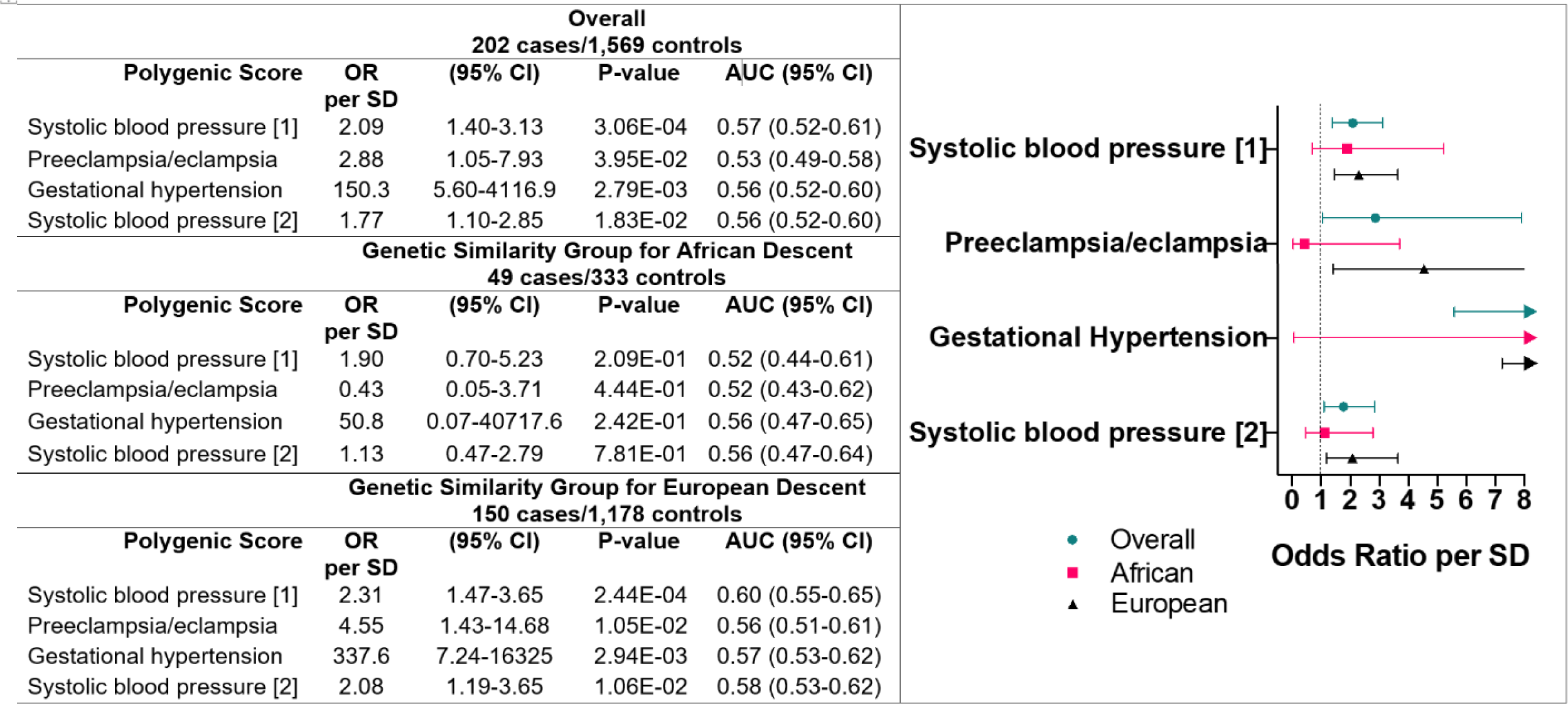
Polygenic score summary statistics and forest plot for odds ratio (ORs) per standard deviation (SD) for hypertensive disorders of pregnancy in multi-ancestry models and African and European genetic similarity groups. AUC: area under the curve

For essential hypertension, the two SBP PGS were strongly associated with HDP in both the overall and stratified analyses (**Figure S4**). The pseudo-R^2^ for SBP PGS [2] was the highest at 5.6% (**Table S7**). The receiver operator characteristic curves indicate that SBP PGS [2] is the most predictive of essential hypertension in PEGS (**Figure S7**), evidenced by the clear linear trend in the percentile plot (**Figure S8**).

## Discussion

Our GWAS is the first to identify and validate genetic variants that are significantly associated with HDP in the maternal genome in *RARB* on chromosome 3. By leveraging a multi-ancestry study design, we increased statistical power for genetic discovery, which enabled us to confirm our findings in the clinically phenotyped UKBB cohort.

The two genome-wide significant loci in PEGS were also significantly associated with HDP in the gene-level analysis, where *RARB* and *RN7SL283P* were the top two genes. Retinoic acid receptors are essential for healthy placental and fetal development. The receptor binds retinoic acid, which is the biologically active form of vitamin A that facilitates cellular signaling in embryonic morphogenesis, cell proliferation, differentiation, and apoptosis^45–47^. Maternal retinol is transferred across the placenta to embryos^47^. All-trans-retinoic acid plays a significant role in maintaining immune balance in some tissues and is linked to the etiology of PE, where greater dysregulation in retinoic acid levels in the decidua (maternal endometrium) is observed in PE cases compared to those with a normal pregnancy^48^. A link has been suggested between decidualization and the anti-angiogenic protein soluble fms-like tyrosine kinase-1 (sFLT1), with retinoic acid directly reducing sFLT1 production in the decidua, which affects early vascular development^48^. Furthermore, in GWAS in a Peruvian cohort focused on maternal and offspring genetic effects on PE, rs4241542, a variant that is 80 kb from the lead variant rs61176331, was associated with proteinuria in offspring (*P*=8.48 − 10^−6^)^49^, further supporting the biological validity of this region in *RARB* for HDP. Future research is needed to investigate the interaction at the maternal-fetal interface, including the influence of gene-gene interaction in placental pathophysiology.

While the variant near *LRP1B* was not validated in UKBB, rs114954125 is near *RN7SL283P,* which is a pseudogene. *LRP1B* belongs to the LDL receptor family and is a tumor suppressor gene^50^. In a previous candidate SNP study, the T-allele variant in *LRP1B*, rs35821928, was associated with increased maternal height and decreased maternal weight^51^.

While the MAGMA gene-set analysis did not yield pathways that met the stringent FDR-corrected significance threshold, the top ten most significant gene sets provide intriguing insights into the probable molecular mechanisms of HDP. The pentose phosphate pathway is among these notable gene sets. This metabolic pathway is involved in cellular redox homeostasis, which is related to oxidative stress as a factor in HDP^11,12^. The identification of immune-response pathways and conditions such as autoimmune thyroid disease, graft-versus-host disease, and allograft rejection points to an immunological dysregulation component of HDP. This is consistent with a growing body of research focused on the immunological basis of HDP^52,19,22,48^.

The gene-level analysis revealed a strong peak of genes at chromosome 6 that did not meet the gene-wide significance threshold. *HLA-DRB5* and *BTNL2* belong to the human major histocompatibility (HLA) complex, where they are involved in the generation of an immune response. Additionally, the adipocytokine signaling pathway connects to signaling molecules originating from adipose tissue, which has been linked to various aspects of cardiovascular health. This could indicate adipose tissue malfunction plays a role in HDP and other cardiometabolic disorders of pregnancy such as gestational diabetes^53^. These suggestive findings related to immune response, inflammation, and metabolic dysfunction call for further analysis and interpretation in large pregnancy cohorts.

The differing patterns of suggestive associations in the GWAS of the composite HDP outcome, GH, and PE in PEGS may indicate differences in etiology (**Figure S3**). The heterogeneous nature of HDP has led to the understanding that there are several distinct PE subtypes. A two-stage placental model related to the development of PE^19,54^ demonstrated that an early-onset, more severe subtype may be associated with underlying genetic or environmental factors that result in abnormal placentation. In contrast, a later-onset, less severe subtype may be a consequence of factors such as obesity, diabetes, cardiovascular disorders, or multi-fetal pregnancy^1^. Further, risk factors for GH and PE vary^55^, supporting evidence for differing etiology across the collection of hypertensive disorders. However, a holistic assessment of HDP that combined PE with other maternal hypertensive disorders as a phenotype for GWAS yielded novel, overlapping risk loci identified in models of PE only and PE or other HDP^56^.

Considering polygenic prediction, using a blood pressure PGS to predict HDP has been found to be more effective than using a PE PGS for women who are non-hypertensive before pregnancy^57^. In this study, we assessed the predictive utility of four PGS in the context of HDP and essential hypertension. While these scores were developed in the White British sub-cohort of UKBB^28,44^, our results demonstrate that an SBP PGS is a strong predictor of HDP. In the stratified results, none of the scores exhibited predictive value for HDP for the AFR genetic similarity group, and only the SBP scores were significantly associated with essential hypertension. These findings highlight the need for customizing PGS development to be inclusive of varying genetic similarity backgrounds to improve the accuracy of HDP risk assessment, as the study’s effectiveness in predicting HDP using a PE PGS or GH PGS does not apply to all populations. Furthermore, while the SBP PGS showed promise for HDP prediction and prevention, it is worth investigating how these genetic scores could be combined with other clinical and environmental factors to develop a more holistic approach to HDP prediction and prevention.

This study has several limitations. First, a composite outcome like HDP can obscure disparate etiologies, for example, between PE and GH. Because of the limited sample size, we grouped the heterogeneous conditions that comprise HDP to increase power to detect genetic variant associations. Second, the results are based on data from a particular time point for each participant. The lack of longitudinal data does not allow us to account for the continuum of risk due to factors such as maternal age and gestational age at the onset of HDP. Further, the study design does not consider the recurrence of complications in subsequent pregnancies. Third, the outcomes are dichotomized due to the nature of the questionnaires completed, which may result in the oversimplification of adverse pregnancy outcomes. Fourth, misclassification bias may be present, since participants who enrolled in PEGS before a pregnancy complication occurred were included in the control group. Fourth, there is a lack of harmonization of phenotypes between study cohorts. While PEGS data are limited, the UKBB uses a wide variety of diagnosis codes for phenotyping, and thus not all pregnancy-related adverse outcomes were excluded from the control group.

## Perspectives (Conclusion)

Variants near *RARB* were significantly associated with HDP in a multi-ancestry cohort. We validated this association in a clinically phenotyped cohort. In gene-set analyses, probable mechanisms associated with HDP included inflammatory and immunological pathways. These findings are supported by recent research focused on the relationship between retinoic acid dysregulation and PE. In-depth functional genomics investigations can elucidate the molecular pathways by which genetic variations near *RARB* contribute to the etiology of HDP, such as the influence of these variants on *RARB* expression, epigenetic alterations, and downstream signaling pathways within the placenta and maternal vascular system. The study demonstrates the strong predictive value of SBP PGS for HDP, but further work should investigate approaches to improve the accuracy and inclusivity of risk assessment for HDP.

HDP is a complex and challenging area of obstetrics, with a significant, disproportionate impact on maternal and fetal health across populations. While we focus on the genetic contribution to HDP, examining the role of social determinants of health is also important, in part because environmental factors can influence genetic risk and the development of HDP. Addressing social, lifestyle, and genetic factors is imperative to reduce the burden of HDP and improve maternal and fetal outcomes. Further research in large, diverse pregnancy cohorts is needed to fully understand the underlying mechanisms and develop effective preventative and therapeutic strategies.

### Novelty and Relevance

#### What is new?

- Genetic studies of hypertensive disorders of pregnancy (HDP) have predominantly focused on individuals of European ancestry.
- Our multi-ancestry genome-wide association study is the first to identify and validate genetic variants that are significantly associated with HDP in the maternal genome in the *RARB* gene.

#### What is relevant?

- We identified two novel genome-wide significant associations with HDP in the maternal genome, namely rs114954125 on chromosome 2 and rs61176331 on chromosome 3. The variant rs61176331 was validated in the UK Biobank. Gene-set analyses revealed a suggestive association between HDP and inflammatory/immunological pathways such as the pentose phosphate pathway and autoimmune thyroid disease.

#### Clinical/Pathophysiological Implications?

- Retinoic acid, which binds to *RARB,* inhibits the expression of sFLT1 in decidua.

The effects of genetic variants near RARB on the pathophysiology of HDP could provide insights into the underlying molecular mechanisms.

- Existing genetic scores for SBP, PE, and GH were predictive of HDP and essential hypertension in the overall PEGS cohort but were not predictive of HDP for those of predominantly African ancestry. Future directions include the development of genetic risk prediction modeling inclusive of large, ancestrally diverse populations to improve utility.

## Data Availability

The data used in this study are available to researchers through application to dbGAP 152 the UK Biobank (https://www.ukbiobank.ac.uk/register-apply/). Reference panel data 153 from the 1000 Genomes Project and polygenic scores from the PGS Catalog are 154 publicly available. The summary statistics that support the study findings are available from the corresponding author upon reasonable request.

## Non-standard Abbreviations and Acronyms

AFR: African ancestry
AMR: Admixed American ancestry
EAF: Effect allele frequency
EAS: East Asian ancestry
EUR: European ancestry
GH: Gestational hypertension
GWAS: Genome-wide association study
HDP: Hypertensive disorders of pregnancy
KEGG: Kyoto Encyclopedia of Genes and Genomes
MAC: Minor allele count
MIM: Mendelian Inheritance in Man
PCA: Principal components analysis
PE: Preeclampsia
PEGS: Personalized Environment and Genes Study
PGS: Polygenic score
SAS: South Asian ancestry
SNP: Single nucleotide polymorphism
SNV: Single nucleotide variant
UKBB: UK Biobank

## Acknowledgements

We would like to acknowledge the participants in the PEGS and UK Biobank cohorts for making this work possible. We would also like to thank Hannah Collins Cakar for assistance with manuscript preparation.

## Supplemental Material

Supplemental Methods

Tables S1–S7

Figures S1-S8

References #58-#90

## References

1. Hutcheon JA, Lisonkova S, Joseph KS. Epidemiology of pre-eclampsia and the other hypertensive disorders of pregnancy. Best Pract. Res. Clin. Obstet. Gynaecol. 2011;25:391–403.

2. Khan KS, Wojdyla D, Say L, Gülmezoglu AM, Van Look PF. WHO analysis of causes of maternal death: a systematic review. Lancet Lond. Engl. 2006;367:1066–1074.

3. Sole KB, Staff AC, Räisänen S, Laine K. Substantial decrease in preeclampsia prevalence and risk over two decades: A population-based study of 1,153,227 deliveries in Norway. Pregnancy Hypertens. 2022;28:21–27.

4. Wang W, Xie X, Yuan T, Wang Y, Zhao F, Zhou Z, Zhang H. Epidemiological trends of maternal hypertensive disorders of pregnancy at the global, regional, and national levels: a population-based study. BMC Pregnancy Childbirth. 2021;21:364.

5. Ghulmiyyah L, Sibai B. Maternal mortality from preeclampsia/eclampsia. Semin. Perinatol. 2012;36:56–59.

6. GBD 2015 Maternal Mortality Collaborators. Global, regional, and national levels of maternal mortality, 1990-2015: a systematic analysis for the Global Burden of Disease Study 2015. Lancet. 2016;388:1775–1812.

7. Fasanya HO, Hsiao CJ, Armstrong-Sylvester KR, Beal SG. A critical review on the use of race in understanding racial disparities in preeclampsia. J. Appl. Lab. Med. 2021;6:247–256.

8. Johnson JD, Louis JM. Does race or ethnicity play a role in the origin, pathophysiology, and outcomes of preeclampsia? An expert review of the literature. Am. J. Obstet. Gynecol. 2022;226:S876–S885.

9. Bartsch E, Medcalf KE, Park AL, Ray JG. Clinical risk factors for pre-eclampsia determined in early pregnancy: systematic review and meta-analysis of large cohort studies. BMJ. 2016;353:i1753.

10. Braunthal S, Brateanu A. Hypertension in pregnancy: Pathophysiology and treatment. SAGE Open Med. 2019;7:2050312119843700.

11. Gilbert JS, Ryan MJ, LaMarca BB, Sedeek M, Murphy SR, Granger JP. Pathophysiology of hypertension during preeclampsia: linking placental ischemia with endothelial dysfunction. Am. J. Physiol.-Heart Circ. Physiol. 2008;294:H541–H550.

12. LaMarca B. Endothelial dysfunction. An important mediator in the pathophysiology of hypertension during preeclampsia. Minerva Ginecol. 2012;64:309–320.

13. Braunthal S, Brateanu A. Hypertension in pregnancy: pathophysiology and treatment. SAGE Open Med. 2019;7:2050312119843700.

14. Lyall F, Bulmer JN, Kelly H, Duffie E, Robson SC. Human trophoblast invasion and spiral artery transformation. Am. J. Pathol. 1999;154:1105–1114.

15. Roberts JM, Rich-Edwards JW, McElrath TF, Garmire L, Myatt L, Global Pregnancy Collaboration. Subtypes of preeclampsia: recognition and determining clinical usefulness. Hypertension. 2021;77:1430–1441.

16. Lu H-Q, Hu R. The role of immunity in the pathogenesis and development of pre-eclampsia. Scand. J. Immunol. 2019;90:e12756.

17. Staff AC, Fjeldstad HE, Fosheim IK, Moe K, Turowski G, Johnsen GM, Alnaes-Katjavivi P, Sugulle M. Failure of physiological transformation and spiral artery atherosis: their roles in preeclampsia. Am. J. Obstet. Gynecol. 2022;226:S895–S906.

18. Harmon AC, Cornelius DC, Amaral LM, Faulkner JL, Cunningham MW Jr, Wallace K, LaMarca B. The role of inflammation in the pathology of preeclampsia. Clin. Sci. 2016;130:409–419.

19. LaMarca B. The role of immune activation in contributing to vascular dysfunction and the pathophysiology of hypertension during preeclampsia. Minerva Ginecol. 2010;62:105–120.

20. Aneman I, Pienaar D, Suvakov S, Simic TP, Garovic VD, McClements L. Mechanisms of key innate immune cells in early- and late-onset preeclampsia. Front. Immunol. 2020;11:1864.

21. Smith SD, Dunk CE, Aplin JD, Harris LK, Jones RL. Evidence for immune cell involvement in decidual spiral arteriole remodeling in early human pregnancy. Am. J. Pathol. 2009;174:1959–1971.

22. LaMarca B, Cornelius D, Wallace K. Elucidating immune mechanisms causing hypertension during pregnancy. Physiology. 2013;28:225–233.

23. Gray KJ, Kovacheva VP, Mirzakhani H, Bjonnes AC, Almoguera B, DeWan AT, Triche EW, Saftlas AF, Hoh J, Bodian DL, et al. Gene-centric analysis of preeclampsia identifies maternal association at PLEKHG1. Hypertension. 2018;72:408–416.

24. McGinnis R, Steinthorsdottir V, Williams NO, Thorleifsson G, Shooter S, Hjartardottir S, Bumpstead S, Stefansdottir L, Hildyard L, Sigurdsson JK, et al. Variants in the fetal genome near FLT1 are associated with risk of preeclampsia. Nat. Genet. 2017;49:1255–1260.

25. Bycroft C, Freeman C, Petkova D, Band G, Elliott LT, Sharp K, Motyer A, Vukcevic D, Delaneau O, O’Connell J, et al. The UK Biobank resource with deep phenotyping and genomic data. Nature. 2018;562:203–209.

26. Mills MC, Rahal C. A scientometric review of genome-wide association studies. Commun. Biol. 2019;2:1–11.

27. Peterson RE, Kuchenbaecker K, Walters RK, Chen C-Y, Popejoy AB, Periyasamy S, Lam M, Iyegbe C, Strawbridge RJ, Brick L, et al. Genome-wide association studies in ancestrally diverse populations: opportunities, methods, pitfalls, and recommendations. Cell. 2019;179:589–603.

28. Honigberg MC, Truong B, Khan RR, Xiao B, Bhatta L, Vy HMT, Guerrero RF, Schuermans A, Selvaraj MS, Patel AP, et al. Polygenic prediction of preeclampsia and gestational hypertension. Nat. Med. 2023;29:1540–1549.

29. Pan UKBB | Pan UKBB [Internet]. [cited 2021 Mar 30];Available from: https://pan.ukbb.broadinstitute.org/

30. National Institute of Environmental Health Sciences (NIEHS). Personalized Environment and Genes Study [Internet]. clinicaltrials.gov; 2022 [cited 2022 Sep 1]. Available from: https://clinicaltrials.gov/ct2/show/NCT00341237

31. Manichaikul A, Mychaleckyj JC, Rich SS, Daly K, Sale M, Chen W-M. Robust relationship inference in genome-wide association studies. Bioinformatics. 2010;26:2867–2873.

32. Maples BK, Gravel S, Kenny EE, Bustamante CD. RFMix: a discriminative modeling approach for rapid and robust local-ancestry inference. Am. J. Hum. Genet. 2013;93:278–288.

33. Diaz-Papkovich A, Anderson-Trocmé L, Ben-Eghan C, Gravel S. UMAP reveals cryptic population structure and phenotype heterogeneity in large genomic cohorts. PLOS Genet. 2019;15:e1008432.

34. : Return 2442 [Internet]. [cited 2023 Jul 19];Available from: https://biobank.ndph.ox.ac.uk/ukb/dset.cgi?id=2442

35. Chang CC, Chow CC, Tellier LC, Vattikuti S, Purcell SM, Lee JJ. Second-generation PLINK: rising to the challenge of larger and richer datasets. GigaScience [Internet]. 2015 [cited 2021 Aug 30];4. Available from: 10.1186/s13742-015-0047-8

36. Firth D. Bias reduction of maximum likelihood estimates. Biometrika. 1993;80:27–38.

37. Yang J, Ferreira T, Morris AP, Medland SE, Genetic Investigation of ANthropometric Traits (GIANT) Consortium, DIAbetes Genetics Replication And Meta-analysis (DIAGRAM) Consortium, Madden PAF, Heath AC, Martin NG, Montgomery GW, et al. Conditional and joint multiple-SNP analysis of GWAS summary statistics identifies additional variants influencing complex traits. Nat. Genet. 2012;44:369–375, S1-3.

38. Willer CJ, Li Y, Abecasis GR. METAL: fast and efficient meta-analysis of genomewide association scans. Bioinformatics. 2010;26:2190–2191.

39. Aken BL, Ayling S, Barrell D, Clarke L, Curwen V, Fairley S, Fernandez Banet J, Billis K, García Girón C, Hourlier T, et al. The Ensembl gene annotation system. Database J. Biol. Databases Curation. 2016;2016:baw093.

40. de Leeuw CA, Mooij JM, Heskes T, Posthuma D. MAGMA: generalized gene-set analysis of GWAS data. PLoS Comput. Biol. 2015;11:e1004219.

41. Zhang S. A simplified protocol for performing MAGMA/H-MAGMA gene set analysis utilizing high-performance computing environments. STAR Protoc. 2022;3:101083.

42. Kanehisa M, Goto S. KEGG: Kyoto Encyclopedia of Genes and Genomes. Nucleic Acids Res. 2000;28:27–30.

43. Kanehisa M, Furumichi M, Sato Y, Ishiguro-Watanabe M, Tanabe M. KEGG: integrating viruses and cellular organisms. Nucleic Acids Res. 2021;49:D545–D551.

44. Kauko A, Aittokallio J, Vaura F, Ji H, Ebinger JE, Niiranen T, Cheng S. Sex differences in genetic risk for hypertension. Hypertension. 2021;78:1153–1155.

45. Huebner H, Hartner A, Rascher W, Strick RR, Kehl S, Heindl F, Wachter DL, Beckmann MW, Fahlbusch FB, Ruebner M. Expression and regulation of retinoic acid receptor responders in the human placenta. Reprod. Sci. 2018;25:1357–1370.

46. RARB retinoic acid receptor beta [Homo sapiens (human)] - Gene - NCBI [Internet]. [cited 2022 Sep 24];Available from: https://www.ncbi.nlm.nih.gov/gene/5915#summary

47. Rhinn M, Dollé P. Retinoic acid signalling during development. Development. 2012;139:843–858.

48. Rajakumar A, Kane MA, Yu J, Taylor RN, Sidell N. Aberrant retinoic acid production in the decidua: Implications for pre-eclampsia. J. Obstet. Gynaecol. Res. 2020;46:1007–1016.

49. Nieves-Colón MA, Badillo Rivera KM, Sandoval K, Villanueva Dávalos V, Enriquez Lencinas LE, Mendoza-Revilla J, Adhikari K, González-Buenfil R, Chen JW, Zhang ET, et al. Clotting factor genes are associated with preeclampsia in high-altitude pregnant women in the Peruvian Andes. Am. J. Hum. Genet. 2022;109:1117–1139.

50. Liu X, White S, Peng B, Johnson AD, Brody JA, Li AH, Huang Z, Carroll A, Wei P, Gibbs R, et al. WGSA: an annotation pipeline for human genome sequencing studies. J. Med. Genet. 2016;53:111–112.

51. Løset M, Johnson MP, Melton PE, Ang W, Huang R-C, Mori TA, Beilin LJ, Pennell C, Roten LT, Iversen A-C, et al. Preeclampsia and cardiovascular disease share genetic risk factors on chromosome 2q22. Pregnancy Hypertens. Int. J. Womens Cardiovasc. Health. 2014;4:178–185.

52. LaMarca B. Endothelial dysfunction; an important mediator in the Pathophysiology of Hypertension during Preeclampsia. Minerva Ginecol. 2012;64:309–320.

53. Trivett C, Lees ZJ, Freeman DJ. Adipose tissue function in healthy pregnancy, gestational diabetes mellitus and pre-eclampsia. Eur. J. Clin. Nutr. 2021;75:1745–1756.

54. Staff AC. The two-stage placental model of preeclampsia: An update. J. Reprod. Immunol. 2019;134–135:1–10.

55. Li X, Tan H, Huang X, Zhou S, Hu S, Wang X, Xu X, Liu Q, Wen SW. Similarities and differences between the risk factors for gestational hypertension and preeclampsia: A population based cohort study in south China. Pregnancy Hypertens. 2016;6:66–71.

56. Tyrmi JS, Kaartokallio T, Lokki AI, Jääskeläinen T, Kortelainen E, Ruotsalainen S, Karjalainen J, Ripatti S, Kivioja A, Laisk T, et al. Genetic risk factors associated with preeclampsia and hypertensive disorders of pregnancy. JAMA Cardiol. 2023;8:674–683.

57. Nurkkala J, Kauko A, FinnGen, Laivuori H, Saarela T, Tyrmi JS, Vaura F, Cheng S, Bello NA, Aittokallio J, et al. Associations of polygenic risk scores for preeclampsia and blood pressure with hypertensive disorders of pregnancy. J. Hypertens. 2023;41:380.

58. gatk4-genome-processing-pipeline [Internet]. 2022 [cited 2022 Sep 6];Available from: https://github.com/gatk-workflows/gatk4-genome-processing-pipeline

59. Auwera GAV der, O’Connor BD. Genomics in the cloud: Using Docker, GATK, and WDL in Terra. O’Reilly Media, Inc.; 2020.

60. Li H. Aligning sequence reads, clone sequences and assembly contigs with BWA-MEM [Internet]. 2013 [cited 2022 Sep 6];Available from: http://arxiv.org/abs/1303.3997

61. Picard Tools - By Broad Institute [Internet]. [cited 2022 Sep 6];Available from: https://broadinstitute.github.io/picard/

62. applybqsr — Clara Parabricks v3.7 documentation [Internet]. [cited 2022 Sep 6];Available from: https://docs.nvidia.com/clara/parabricks/3.7.0/Documentation/ToolDocs/man_applybqsr.html

63. DeepVariant [Internet]. 2022 [cited 2022 Sep 6];Available from: https://github.com/google/deepvariant

64. Liu X, White S, Peng B, Johnson AD, Brody JA, Li AH, Huang Z, Carroll A, Wei P, Gibbs R, et al. WGSA: an annotation pipeline for human genome sequencing studies. J. Med. Genet. 2016;53:111–112.

65. Aken BL, Ayling S, Barrell D, Clarke L, Curwen V, Fairley S, Fernandez Banet J, Billis K, García Girón C, Hourlier T, et al. The Ensembl gene annotation system. Database J. Biol. Databases Curation. 2016;2016:baw093.

66. Manichaikul A, Mychaleckyj JC, Rich SS, Daly K, Sale M, Chen W-M. Robust relationship inference in genome-wide association studies. Bioinformatics. 2010;26:2867–2873.

67. Population Structure and Relatedness Inference using the GENESIS Package [Internet]. [cited 2021 Feb 28];Available from: https://www.bioconductor.org/packages/devel/bioc/vignettes/GENESIS/inst/doc/pcair.html

68. Conomos MP, Miller MB, Thornton TA. Robust inference of population structure for ancestry prediction and correction of stratification in the presence of relatedness. Genet. Epidemiol. 2015;39:276–293.

69. Zheng X, Levine D, Shen J, Gogarten SM, Laurie C, Weir BS. A high-performance computing toolset for relatedness and principal component analysis of SNP data. Bioinformatics. 2012;28:3326–3328.

70. Delaneau O, Zagury J-F, Robinson MR, Marchini JL, Dermitzakis ET. Accurate, scalable and integrative haplotype estimation. Nat. Commun. 2019;10:5436.

71. Maples BK, Gravel S, Kenny EE, Bustamante CD. RFMix: a discriminative modeling approach for rapid and robust local-ancestry inference. Am. J. Hum. Genet. 2013;93:278–288.

72. Byrska-Bishop M, Evani US, Zhao X, Basile AO, Abel HJ, Regier AA, Corvelo A, Clarke WE, Musunuri R, Nagulapalli K, et al. High-coverage whole-genome sequencing of the expanded 1000 Genomes Project cohort including 602 trios. Cell. 2022;185:3426–3440.e19.

73. IGSR | samples [Internet]. [cited 2022 May 9];Available from: https://www.internationalgenome.org/data-portal/sample

74. Bycroft C, Freeman C, Petkova D, Band G, Elliott LT, Sharp K, Motyer A, Vukcevic D, Delaneau O, O’Connell J, et al. The UK Biobank resource with deep phenotyping and genomic data. Nature. 2018;562:203–209.

75. : Category 100319 [Internet]. [cited 2022 Sep 9];Available from: https://biobank.ndph.ox.ac.uk/ukb/label.cgi?id=100319

76. : Resource 1671 [Internet]. [cited 2022 Sep 9];Available from: https://biobank.ctsu.ox.ac.uk/crystal/refer.cgi?id=1671

77. : Return 2442 [Internet]. [cited 2023 Jul 19];Available from: https://biobank.ndph.ox.ac.uk/ukb/dset.cgi?id=2442

78. Quality Control (QC) | Pan UKBB [Internet]. [cited 2021 Apr 9];Available from: https://pan-dev.ukbb.broadinstitute.org/docs/qc

79. Chang CC, Chow CC, Tellier LC, Vattikuti S, Purcell SM, Lee JJ. Second-generation PLINK: rising to the challenge of larger and richer datasets. GigaScience [Internet]. 2015 [cited 2021 Aug 30];4. Available from: 10.1186/s13742-015-0047-8

80. Yang J, Ferreira T, Morris AP, Medland SE, Genetic Investigation of ANthropometric Traits (GIANT) Consortium, DIAbetes Genetics Replication And Meta-analysis (DIAGRAM) Consortium, Madden PAF, Heath AC, Martin NG, Montgomery GW, et al. Conditional and joint multiple-SNP analysis of GWAS summary statistics identifies additional variants influencing complex traits. Nat. Genet. 2012;44:369–375, S1-3.

81. Willer CJ, Li Y, Abecasis GR. METAL: fast and efficient meta-analysis of genomewide association scans. Bioinformatics. 2010;26:2190–2191.

82. Durinck S, Moreau Y, Kasprzyk A, Davis S, De Moor B, Brazma A, Huber W. BioMart and Bioconductor: a powerful link between biological databases and microarray data analysis. Bioinforma. Oxf. Engl. 2005;21:3439–3440.

83. Durinck S, Spellman PT, Birney E, Huber W. Mapping identifiers for the integration of genomic datasets with the R/Bioconductor package biomaRt. Nat. Protoc. 2009;4:1184–1191.

84. de Leeuw CA, Mooij JM, Heskes T, Posthuma D. MAGMA: generalized gene-set analysis of GWAS data. PLoS Comput. Biol. 2015;11:e1004219.

85. Zhang S. A simplified protocol for performing MAGMA/H-MAGMA gene set analysis utilizing high-performance computing environments. STAR Protoc. 2022;3:101083.

86. Liberzon A, Birger C, Thorvaldsdóttir H, Ghandi M, Mesirov JP, Tamayo P. The Molecular Signatures Database (MSigDB) hallmark gene set collection. Cell Syst. 2015;1:417–425.

87. Kanehisa M, Goto S. KEGG: Kyoto Encyclopedia of Genes and Genomes. Nucleic Acids Res. 2000;28:27–30.

88. Kanehisa M, Furumichi M, Sato Y, Ishiguro-Watanabe M, Tanabe M. KEGG: integrating viruses and cellular organisms. Nucleic Acids Res. 2021;49:D545–D551.

89. Kauko A, Aittokallio J, Vaura F, Ji H, Ebinger JE, Niiranen T, Cheng S. Sex differences in genetic risk for hypertension. Hypertension. 2021;78:1153–1155.

90. Honigberg MC, Truong B, Khan RR, Xiao B, Bhatta L, Vy HMT, Guerrero RF, Schuermans A, Selvaraj MS, Patel AP, et al. Polygenic prediction of preeclampsia and gestational hypertension. Nat. Med. 2023;29:1540–1549.

